# Bamlanivimab does not neutralize two SARS-CoV-2 variants carrying E484K in vitro

**DOI:** 10.1101/2021.02.24.21252372

**Authors:** Marek Widera, Alexander Wilhelm, Sebastian Hoehl, Christiane Pallas, Niko Kohmer, Timo Wolf, Holger F Rabenau, Victor Corman, Christian Drosten, Maria JGT Vehreschild, Udo Goetsch, Rene Gottschalk, Sandra Ciesek

**Author notes:** Correspondence: Prof. Dr. med. Sandra Ciesek, Institute for Medical. Virology, University Hospital Frankfurt, Paul-Ehrlich-Str.40, 60596 Frankfurt am Main.

## Abstract

The IgG1 monoclonal antibody (mAb) bamlanivimab (LY-CoV555) prevents viral attachment and entry into human cells by blocking attachment to the ACE2 receptor. However, whether bamlanivimab is equally effective against SARS-CoV-2 emerging variants of concern (VOC) is not fully known. Hence, the aim of this study was to determine whether bamlanivimab is equally effective against SARS-CoV-2 emerging VOC. The ability of bamlanivimab to neutralize five SARS-CoV-2 variants including B.1.1.7 (mutations include N501Y and del69/70), B.1.351 (mutations include E484K and N501Y) and P.2 (mutations include E484K in the absence of a N501Y mutation) was analyzed in infectious cell culture using CaCo2 cells. Additionally, we analyzed vaccine-elicited sera after immunization with BNT162b2, and convalescent sera for its ability to neutralize SARS-CoV-2 variants.

We found that the variant B.1.1.7, as well as two isolates from early 2020 (FFM1 and FFM7) could be efficiently neutralized by bamlanivimab (titer 1/1280, respectively), however, no neutralization effect could be detected against either B.1.135 or P.2, both harboring the E484K substitution. Vaccine-elicited sera showed slightly decreased neutralizing activity against B1.1.7, B.1.135 and P.2

Our in vitro findings indicate that, in contrast to vaccine-elicited sera, bamlanivimab may not provide efficacy against SARS-CoV-2 variants harboring the E484K substitution. Confirmation of the SARS-CoV-2 variant, including screening for E484K, may be needed before initiating mAb treatment with bamlanivimab to ensure both efficacious and efficient use of the antibody product. Hence, variant-specific mAb agents may be required to treat emerging VOC.

---

As vaccination campaigns against Covid-19 are ongoing, the majority of the world’s population remains unimmunized, and many at risk for severe disease. The availability of both therapeutic and prophylactic agents with proven efficacy are urgently needed. The IgG1 monoclonal antibody (mAb) bamlanivimab (LY-CoV555) has been demonstrated to accelerate the decline in viral load.(1) The mAb prevents viral attachment and entry into human cells by binding to the receptor-binding domain in the viral spike protein and blocking attachment to ACE2. Recently, bamlanivimab has been authorized by the FDA for emergency use in early mild to moderate COVID-19 disease.

Emerging “variants of concern” SARS-CoV-2 referred to as B.1.1.7, B.1.351, P.1, and P.2 initially observed in the United Kingdom, South Africa, and Brazil, respectively, are in the process of fixation in the population. Mutations in the spike’s receptor binding domain, in particular N501Y, were associated with increased infectivity due to enhanced receptor binding(2). Further studies have shown that of E484 represents an immunodominant site on the RBD since E484K reduced naturalization capacity of human convalescent sera by >100-fold.(3) These amino acid substitutions have also been observed to evade the antibody response elicited by an infection with other SARS-CoV-2 variants, or vaccination.(4) So far, it is not yet known whether bamlanivimab is equally effective against these emerging “variants of concern”.

In this study, we examined the ability of bamlanivimab, vaccine-elicited sera after immunization with BNT162b2, and convalescent sera, to neutralize the emerging SARS-CoV-2 variants B.1.1.7 (mutations include N501Y and Δ69/70), B.1.351 (mutations include E484K and N501Y) and P.2 (mutations include E484K in the absence of a 501 mutation). These isolates have been obtained from travelers from Great Britain, South Africa and Brazil, respectively. Two SARS-CoV-2 isolates collected in early 2020 were also tested (FFM1, FFM7)(5, 6). The mAb working solution was serially diluted 1:2 and incubated with 4000 TCID_50_ / ml of each SARS-CoV-2 isolate, and subjected to Caco2 cell based SARS-CoV-2 neutralization assay. The corresponding sample dilution resulting in 50% virus neutralization titer (NT_50_) was determined. After 3 days of incubation cells were evaluated for the presence of a cytopathic effect (CPE).

The variant B.1.1.7, as well as the isolates from early 2020 (FFM1 and FFM7), could be efficiently neutralized by bamlanivimab (titer 1/1280, respectively) (**table 1**). However, no neutralization effect could be detected against either B.1.135 or P.2, both harboring the E484K substitution. Spike protein alignments confirmed that only E484K substitution occurs exclusively in the two strains that could not be neutralized by bamlanivimab.

**Table 1:**
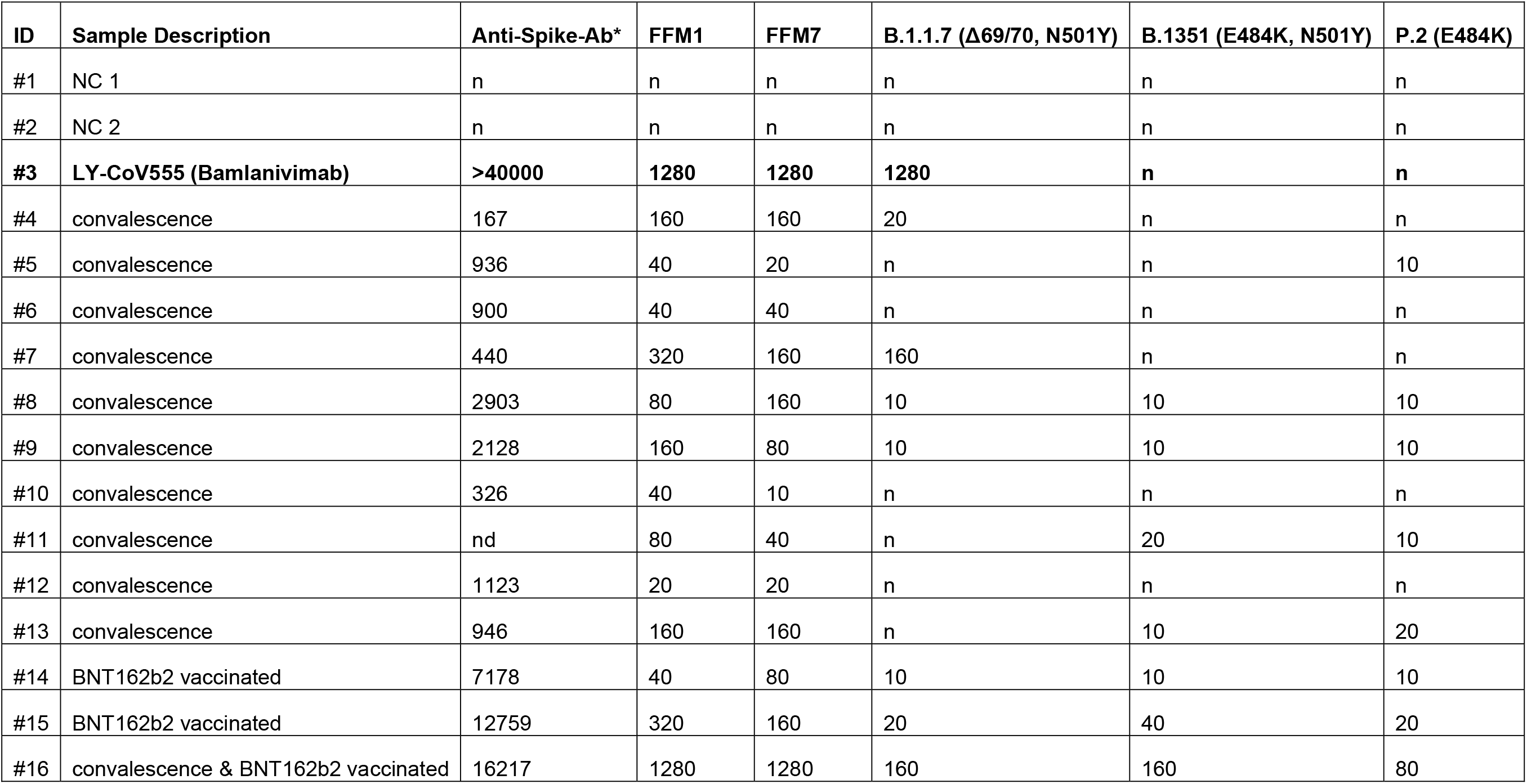
Neutralization titers against SARS-CoV-2 variants. The mAb solution was serially diluted (1:2) and incubated with 4000 TCID_50_ / ml of the indicated SARS-CoV-2 variant. Caco2 cells subsequently inoculated and analyzed for a CPE formation after 72 h incubation. Values indicate SARS-CoV-2 microneutralization titer (NT_50_), resulting in 50% virus neutralization. The values indicate mean values from two replicates per cell line. n= no neutralization. NC = negative control sera. * tested with SARS-CoV-2 IgG II Quant (Abbott Diagnostics, Delkenheim, Germany) using the automated Alinity i device. The quantitative assay is for the detection of - among others - neutralizing antibodies against the receptor binding domain (RBD) of the S1 subunit of the spike protein of SARS-CoV-2. Results are given as AU/mL (analytical measurement range: 21.0 - 40 000.0).

Vaccine-elicited sera showed neutralizing activity against FFM1 and FFM7, and slightly decreased activity against B1.1.7, B.1.135 and P.2 (**table 1**). In one exceptional case of serum from an individual who was vaccinated after convalescence, high titers that were equivalent to the monoclonal antibody preparation were observed. Convalescent sera had lower neutralizing activity against “variants of concern” when compared to the variants from early 2020, which may facilitate re-infection with emerging variants.

Our *in vitr*o findings indicate that, in contrast to vaccine-elicited sera, bamlanivimab may not provide efficacy against SARS-CoV-2 variants harboring the E484K substitution. Our data was in agreement with previous studies using artificial pseudoviruses, showing that LY-CoV555 is ineffective against B.1.351 but is still effective against B.1.1.7 (7, 8). However, this is the first study evaluating the neutralizing capacity of bamlanivimab (LY-CoV555) using live SARS-CoV-2 isolates.

We conclude that confirmation of the SARS-CoV-2 variant, including screening for E484K, may be needed before initiating mAb treatment with bamlanivimab to ensure both efficacious and efficient use of the antibody product. Variant-specific mAb agents may be required to treat emerging “variants of concern”.

## Data Availability

All necessary data are included in the manuscript.

